# Efficacy of physiotherapy in reducing pain in subjects with temporomandibular disorders: a systematic review with network meta-analysis

**DOI:** 10.64898/2026.10.11.26365246

**Authors:** Alessandro Alvisi, Alberto Casalini, Laura Mammana, Alessandro Ugolini

## Abstract

**Objective:** Temporomandibular disorders are common conditions in the general population. Several therapeutic approaches are available, but there is no consensus on the most effective, particularly in physiotherapy. The objective of this review is to assess the effect of manual therapy and therapeutic exercise in reducing pain in patients with temporomandibular disorders.

**Data sources:** Four databases (MEDLINE, PEDro, EMBASE, CINAHL) were searched up to March 2025.

**Review methods:** We included randomized controlled trials evaluating the effects of manual therapy and/or therapeutic exercise in individuals with temporomandibular disorders, compared to any other rehabilitation treatment, with pain intensity as the primary outcome. The methodological quality was assessed using the PEDro scale. A network meta-analysis was performed to compare the effect of different rehabilitative approaches.

**Results:** 25 randomised clinical trials for a total of 1,540 people were included. The main comparisons were: manual therapy and/or therapeutic exercise vs no treatment or placebo, occlusal splint, and instrumental physiotherapy. In most studies, manual therapy and therapeutic exercise were more effective than controls in reducing pain. However, the network meta-analysis indicated that multimodal treatments were the most effective. The methodological quality of the included studies ranged from 3 to 9 points.

**Conclusion:** manual therapy and therapeutic exercise are effective strategies for reducing pain in subjects with temporomandibular disorders and should be used as first-line interventions before more invasive treatments; the combination of dry needling and manual therapy appears to be the most effective approach. Nonetheless, the variability in study quality suggests that further high-quality research is needed to confirm these findings.

## Introduction

Temporomandibular disorders are defined as "a group of musculoskeletal and neuromuscular conditions involving the temporomandibular joints, the masticatory muscles, and all associated tissues"[1]; they are primarily characterized by pain in the jaw or temple area, often accompanied by joint sounds and limited mouth opening, which can impair functions such as chewing, speaking and yawning[2]. Their prevalence in general population ranges between 5% and 12%[3] and affected individuals frequently develop chronic symptoms and comorbid neuromusculoskeletal conditions that negatively impact their quality of life[4].

Treatment aims to reduce pain and improve function, beginning with a conservative, reversible and evidence-based approach[5]. While conservative strategies have shown effect in relieving pain and improving jaw function, the specific role of physiotherapy has been insufficiently explored. Only one prior meta-analysis found no high-quality evidence, reporting mostly small to moderate effect sizes without a clear indication of the superiority of exercise over other conservative treatments and manual therapy demonstrated promising effects[6]. More recent studies have mainly focused on physical modalities such as laser therapy and ultrasound[7].

Given these gaps, an updated analysis of primary studies evaluating physiotherapeutic interventions for temporomandibular disorders is needed. A network meta-analysis is particularly useful and enables indirect comparisons between interventions not directly compared within individual randomised clinical trials; it can enhance the precision of effect estimates and reduce uncertainty about which treatments are most effective and it also facilitates a ranking of therapeutic options based on effect in pain reduction.

## Methods

This systematic review and network meta-analysis was conducted following the Preferred Reporting Items for Systematic Reviews and Meta-Analyses for network meta-analyses extension guidelines [8]. The protocol was registered in the International Prospective Register of Systematic Reviews with number CRD42022347010.

The multimodal approach adopted by many studies included in this review led us to perform a network meta-analysis that helps to identify combined treatments. MEDLINE, EMBASE, PEDro and CINAHL were searched from their inception to March 2025 using a comprehensive strategy (Table 1). Additional records were identified through manual searches. Eligibility criteria were defined as follows:

**Table 1.**
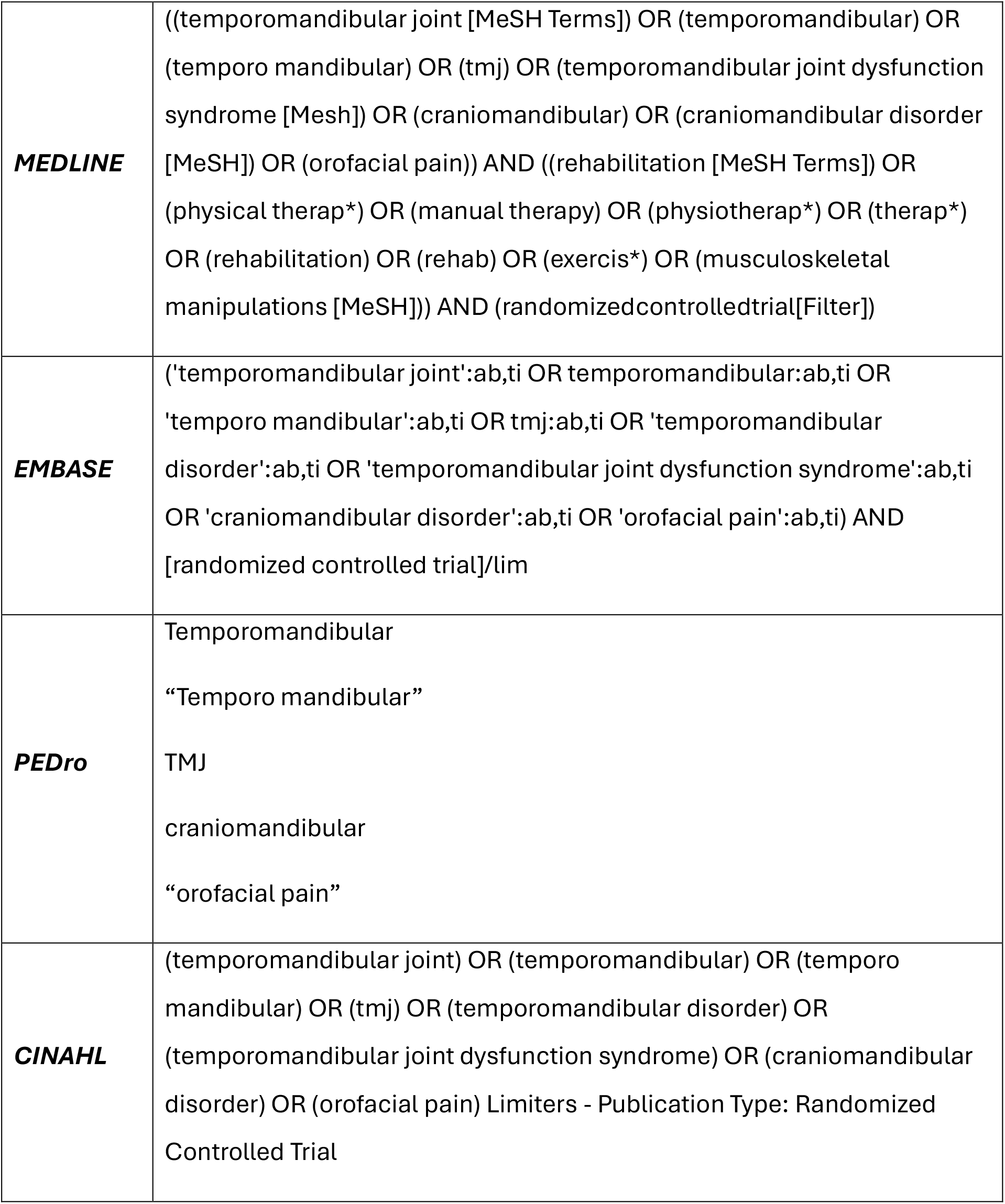
Search strategy used for each database.

- *Participants*: subjects affected by a temporomandibular disorder diagnosed according to the Diagnostic Criteria for Temporomandibular Disorders [9]; people with any pathology other than temporomandibular disorders and those who received oral surgery were excluded.

- *Intervention*: any manual therapy technique or therapeutic exercise applied to any region of the body; interventions involving oral appliances, physical modalities (laser, transcutaneous electrical nerve stimulation, ultrasound) or pharmacological treatments were excluded.

- *Comparison*: any other treatment, including placebo, sham treatments or other techniques. Comparisons with invasive treatments were excluded. When multiple control groups were present, the most competitive comparator was selected for meta-analysis.

- *Outcome*: pain intensity measured using validated tools such as the Visual Analog Scale or Numeric Pain Rating Scale. Only studies reporting numerical outcome measures with variability indices (standard deviation or confidence interval) were included. Studies reporting outcomes only graphically were excluded. If multiple timepoints were available, the first post-treatment value was used for analysis.

- *Study*: randomized clinical trials published in English or Italian.

The screening of the included articles was performed using EndNote® (version X7.0.1).

After the removal of duplicates, two reviewers (AA, LM) independently screened all articles by title, abstract and full text based on the eligibility criteria; any disagreement between them was resolved through discussion and eventually by a third author (AC).

Two independent reviewers (AA, LM) extracted data from the included studies on an electronic spreadsheet (Microsoft Excel® for Microsoft Office 365®); any disagreement between them was resolved through discussion and, eventually, by a third author (AC).

Two reviewers (AA, LM) extracted the following data from the selected articles: title, subjects for each group, treatment used for the intervention and control groups, mean and standard deviation at baseline and post-intervention, treatment duration, outcome measures, duration and number of follow-up(s) and conclusions. For those studies in which education or counseling was adopted both for the intervention and the control group, it was ignored as part of the treatment.

The methodological quality of the included studies was assessed using the PEDro score. For the randomised clinical trials already indexed in PEDro, the score was taken from there, while for the other studies, two authors (AA, LM) independently attributed the score; any disagreement between them was resolved through discussion and eventually by a third author (AC).

Group means, standard deviation and sample sizes were extracted from each study.

When standard deviation was missing or only medians were reported, standard deviation was estimated and medians were converted to means using the method proposed by Wan et al [10]. For effect size between control and experimental groups, Standardized Mean Difference with Hedges’ "g" correction was chosen, as different outcome measures were used across the studies. Two meta-analyses were also conducted, calculating the change score as the effect size to assess whether randomization of studies effectively recruited study samples, comparing it to post-treatment results. The formula reported in the Cochrane Handbook [11] was used, employing a correlation coefficient of 0.8 for standard deviation estimation. Another meta-analysis was conducted with studies considering therapeutic exercise versus manual therapy as control and/or experimental groups to evaluate if one treatment had significant effects compared to the other.

Meta-analysis results were combined using frixed and random-effects models (fixed model results shown in forest plot only) and synthesised into forest plots using RevMan5 formatting [12]. The inverse variance method for study weighting [13] and REML Maximum Likelihood Estimation for tau^2 estimation [13] were used. Statistical heterogeneity among studies was assessed using I², interpreting I² according to established guidelines (0-40%= not important statistical heterogeneity; 30-60%= moderate heterogeneity; 50-75%= substantial heterogeneity; 75-100%= considerable heterogeneity) [12]. Subgroup meta-analyses were conducted to study heterogeneity due to treatment variation across studies [14]. Meta-regressions with bubble plots were performed, using treatment start time, intervention duration and PEDro score as covariates [15]. Publication bias and heterogeneity were investigated using funnel plot [16], radial plot [17], Baujat plot [18], Begg test [19] and Egger test (p>0.05 indicating publication bias risk) [20].

A network meta-analysis was conducted to establish significant differences between therapeutic exercise and manual therapy groups and determine the best treatment combinations. Effect sizes between all pairs were estimated using Mean Difference, then Standardized Mean Difference was used in network meta-analysis to estimate direct and indirect effects, employing random effects for the outcome and a net plot was created to illustrate connections [21]. Heterogeneity components were evaluated within the design and between design inconsistencies, assessed with p<0.05.

To generate a treatment ranking indicating which treatment is more or less likely to produce the greatest benefits, a frequentist approach based on p-scores was used to classify treatments. This measured the certainty that one treatment was better than another, calculated as the average of all competing treatments, where the p-score is equivalent to the Surface Under the Cumulative Ranking (SUCRA) score [22]. The parameter was specified to define if smaller effect sizes (negative) in a comparison indicated a beneficial effect ("good"), as negative effect sizes indicated that a treatment was more effective in pain reduction. A forest plot was produced to visualize uncertainty in the network, evaluating if differences were significant [23] and specifying therapeutic exercise as the reference group. A quadratic heat map was created to visualize design inconsistency, observing changes in the net heat map when assuming a random effects model versus fixed. Finally, a net splitting was conducted to divide estimates between direct and indirect, checking inconsistency in individual comparison estimates within the network (with p<0.05 indicating significant disagreement) between direct and indirect estimates, shown in a forest plot.

Data analysis was performed using R, a free statistical software environment, utilising the "meta" and "netmeta" packages [24].

## Results

### Study selection

The bibliographic research produced 3,663 results (Figure 1); through hand-searching and cross-referencing, five other articles were identified. After removing duplicates, 2,987 articles were screened by title, 392 by abstract and 124 by full text. At the end of the screening process, 25 studies were included in the review [25–49]

**Figure 1.**
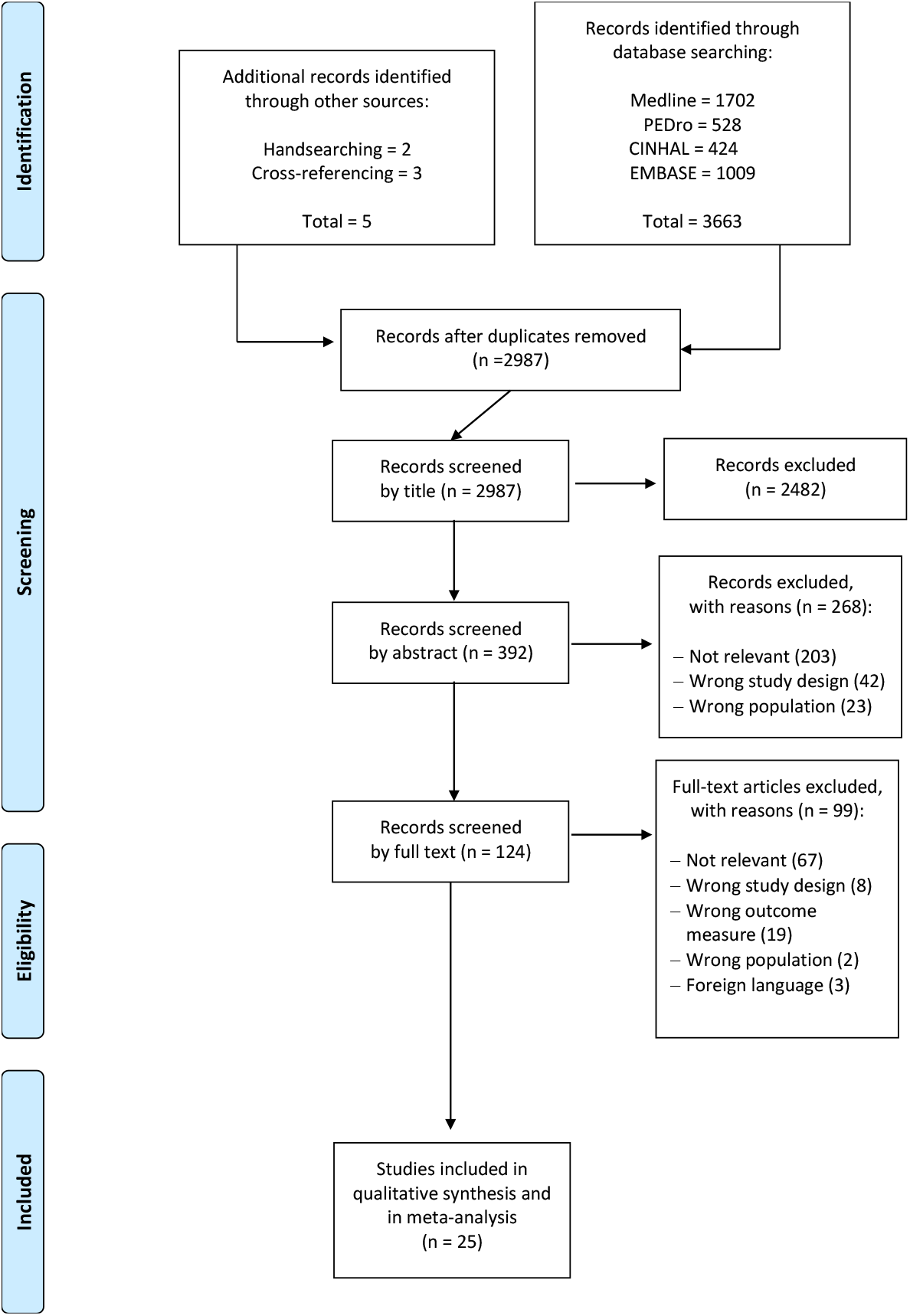
PRISMA flow chart

### Study characteristics

A total of 1,540 subjects were included. Several studies assessed pain intensity through a Visual Analog Scale [25,26,27,29,31,32,34,37,38,39,40,41,42,43,44,45,46,47] or with Numeric Rating Scale [30,33,48,49]. In contrast, others used other validated scores like the 11-point ordinal pain scale [28,36] or the 11-point self-reported graded chronic pain scale [35]. The included studies investigated the effects of different rehabilitative techniques either alone or in combination. The most commonly used instrumental therapies were ultrasound, transcutaneous electrical nerve stimulationand laser therapy. Mean treatment duration was 52 days with a standard deviation of 44,8 days. Follow-up(s), which varied between the studies from one day [37–38] to six months [47].

Main characteristics of each included study are reported in Table 2.

**Table 2.**
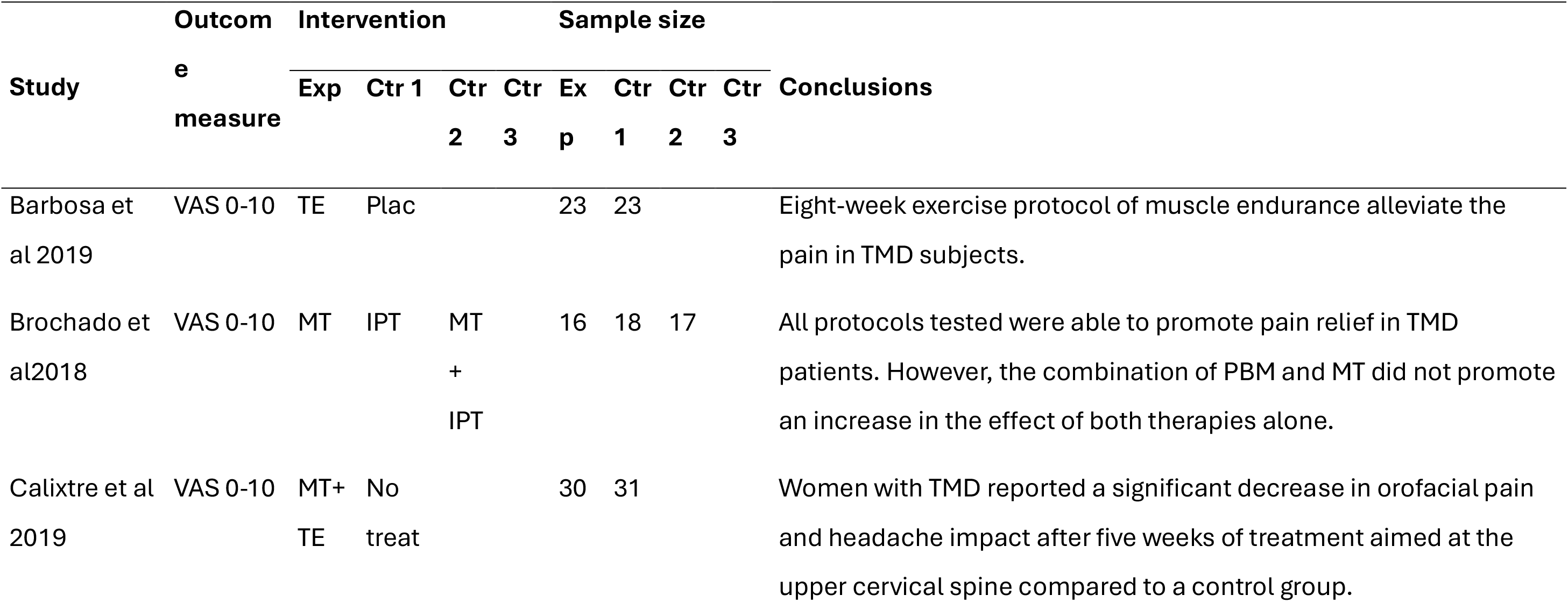
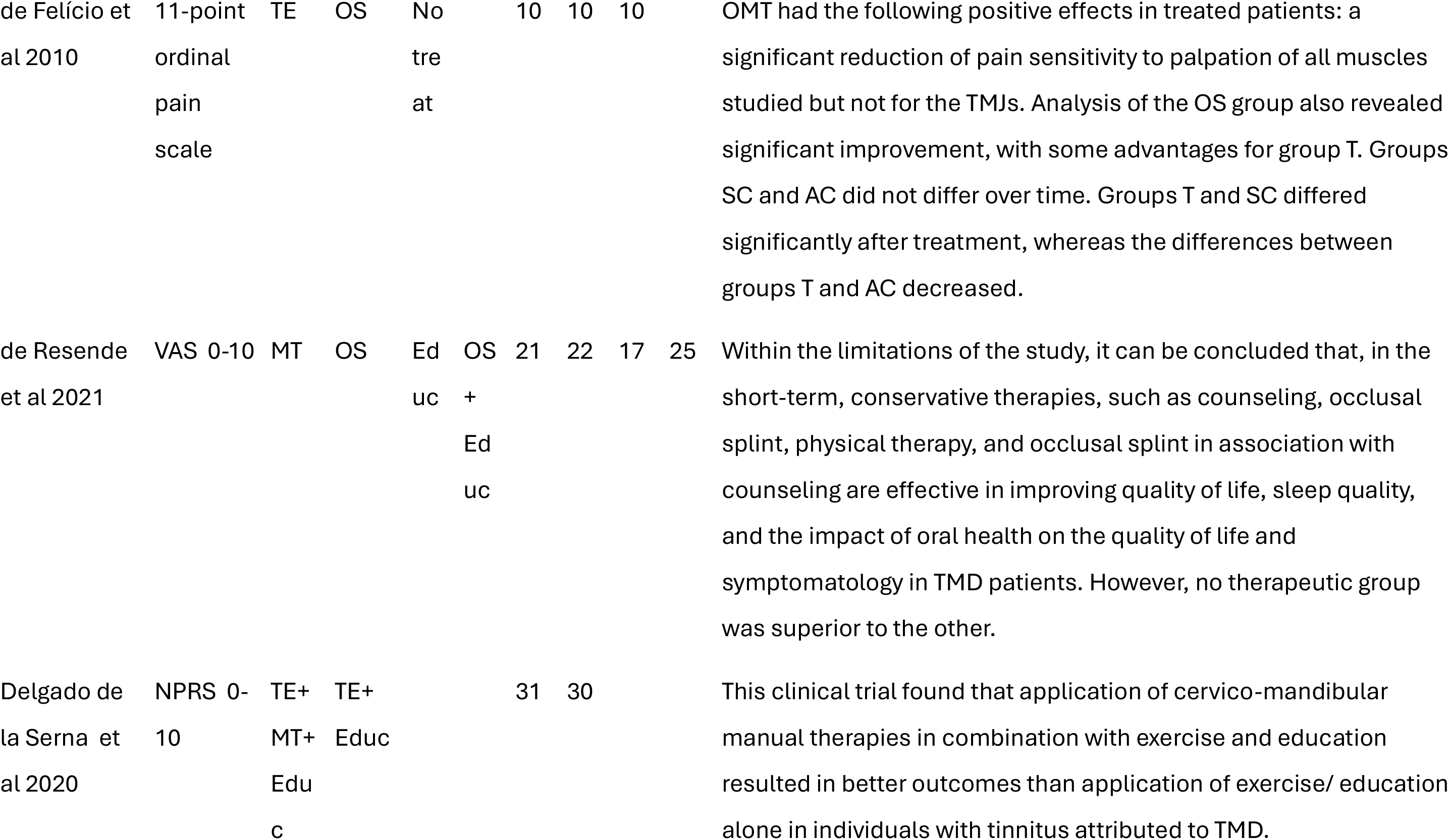
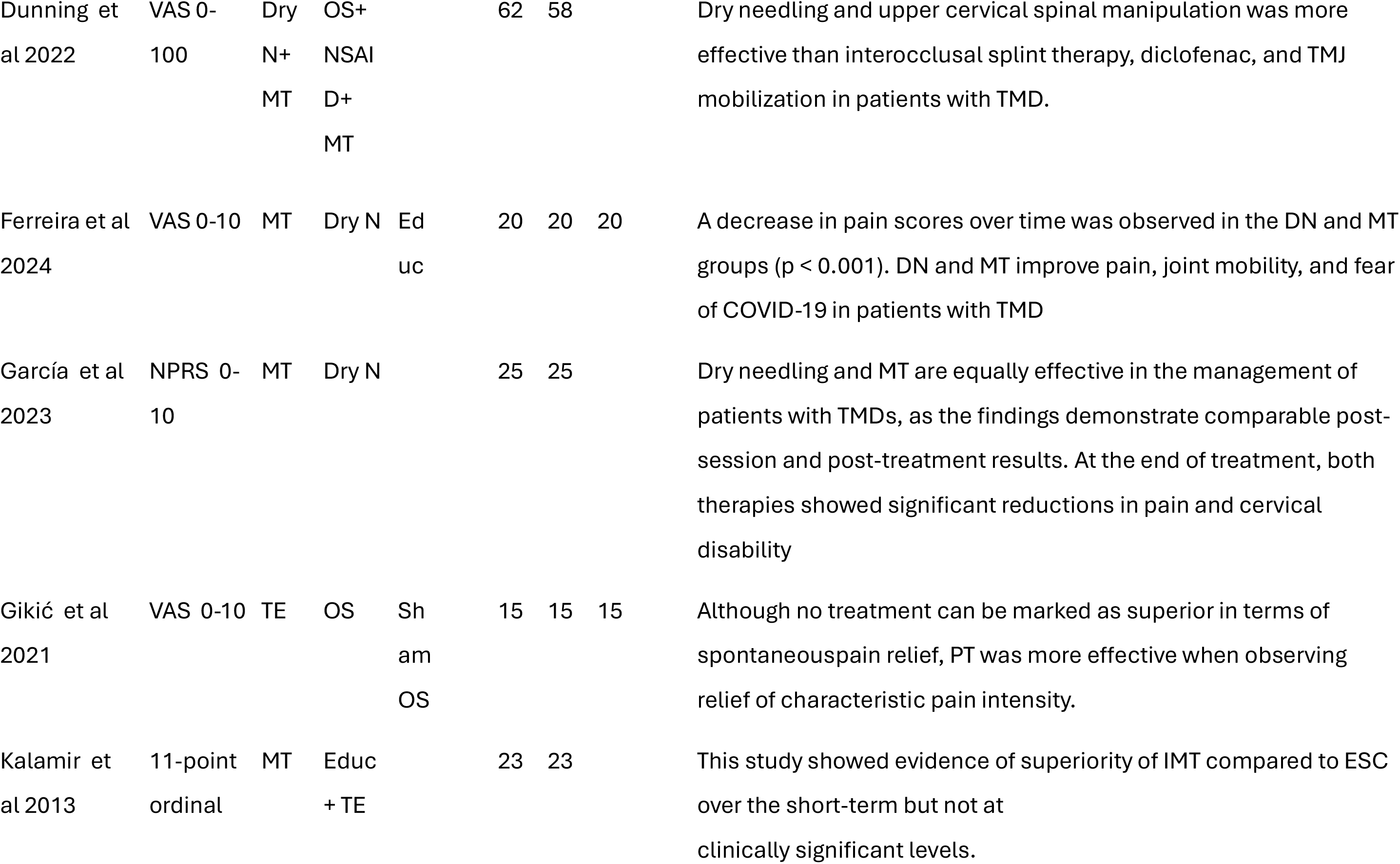
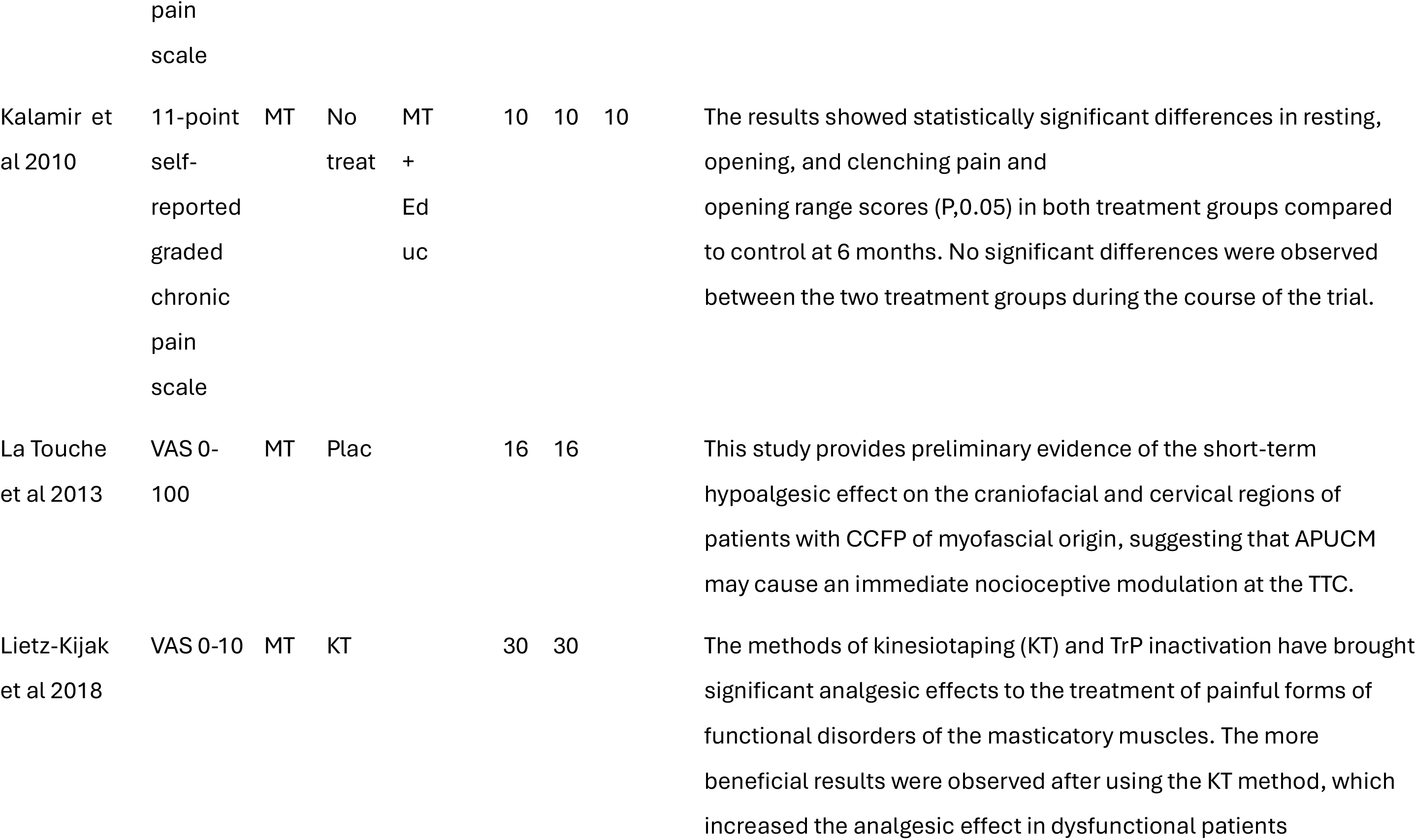
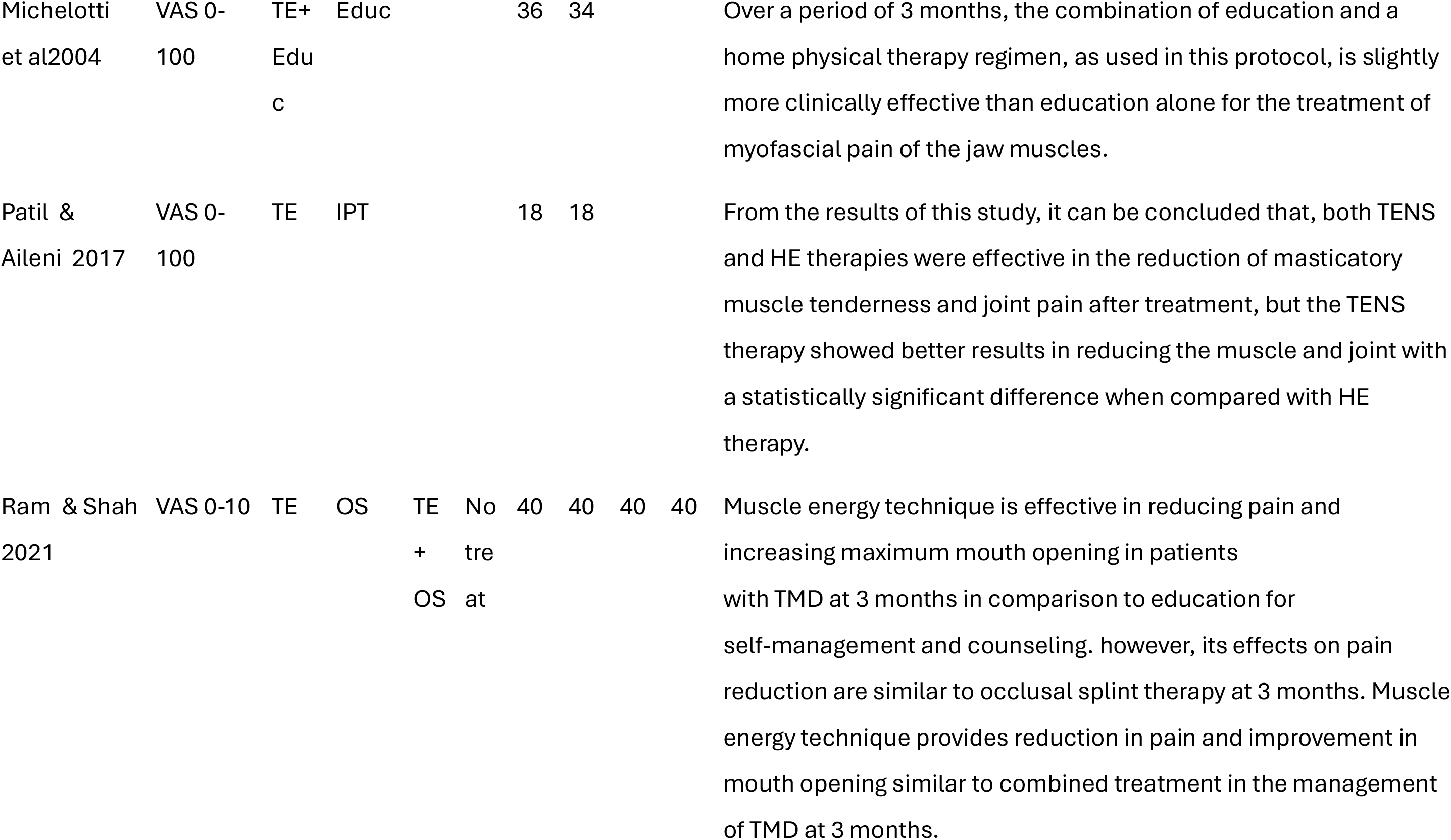
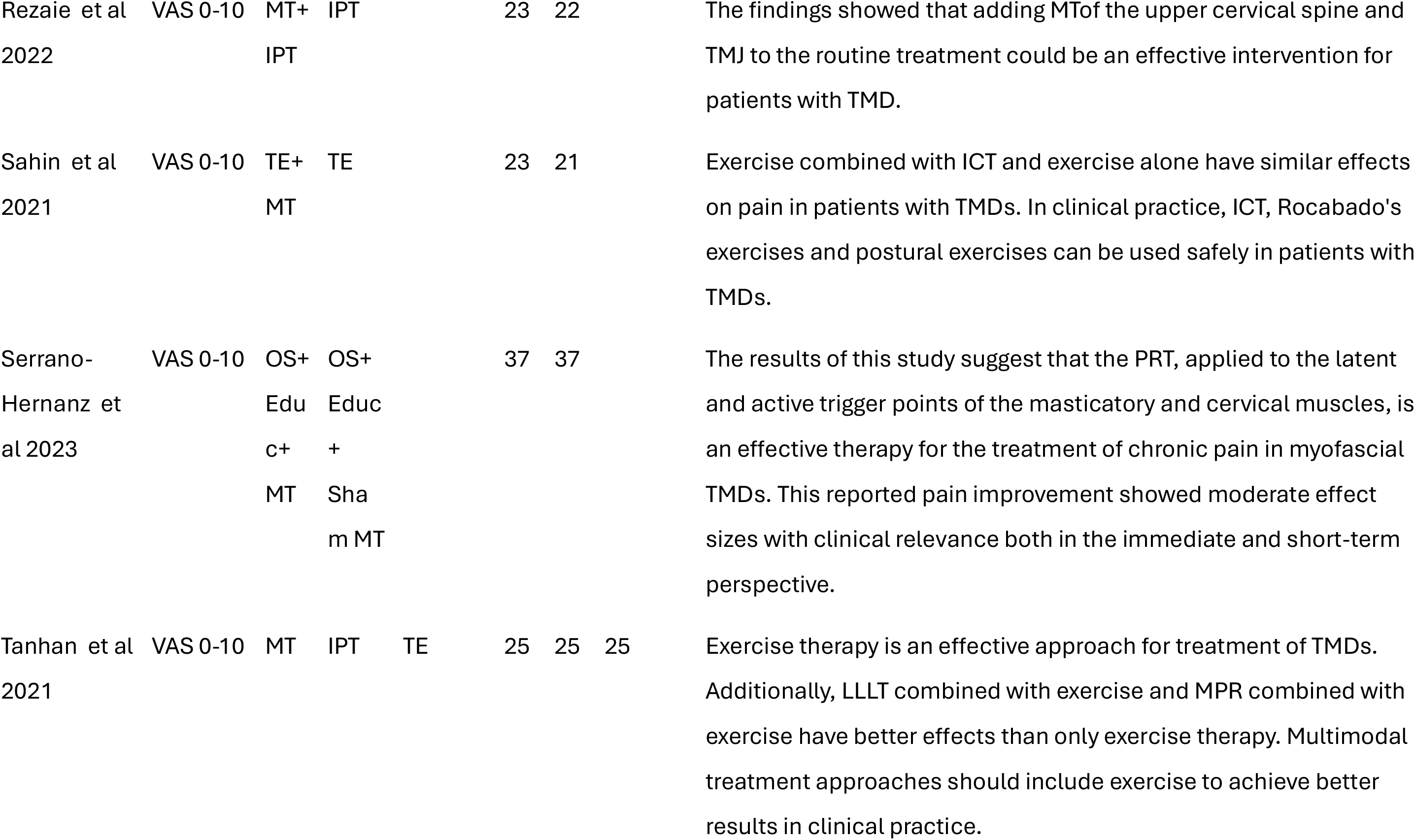
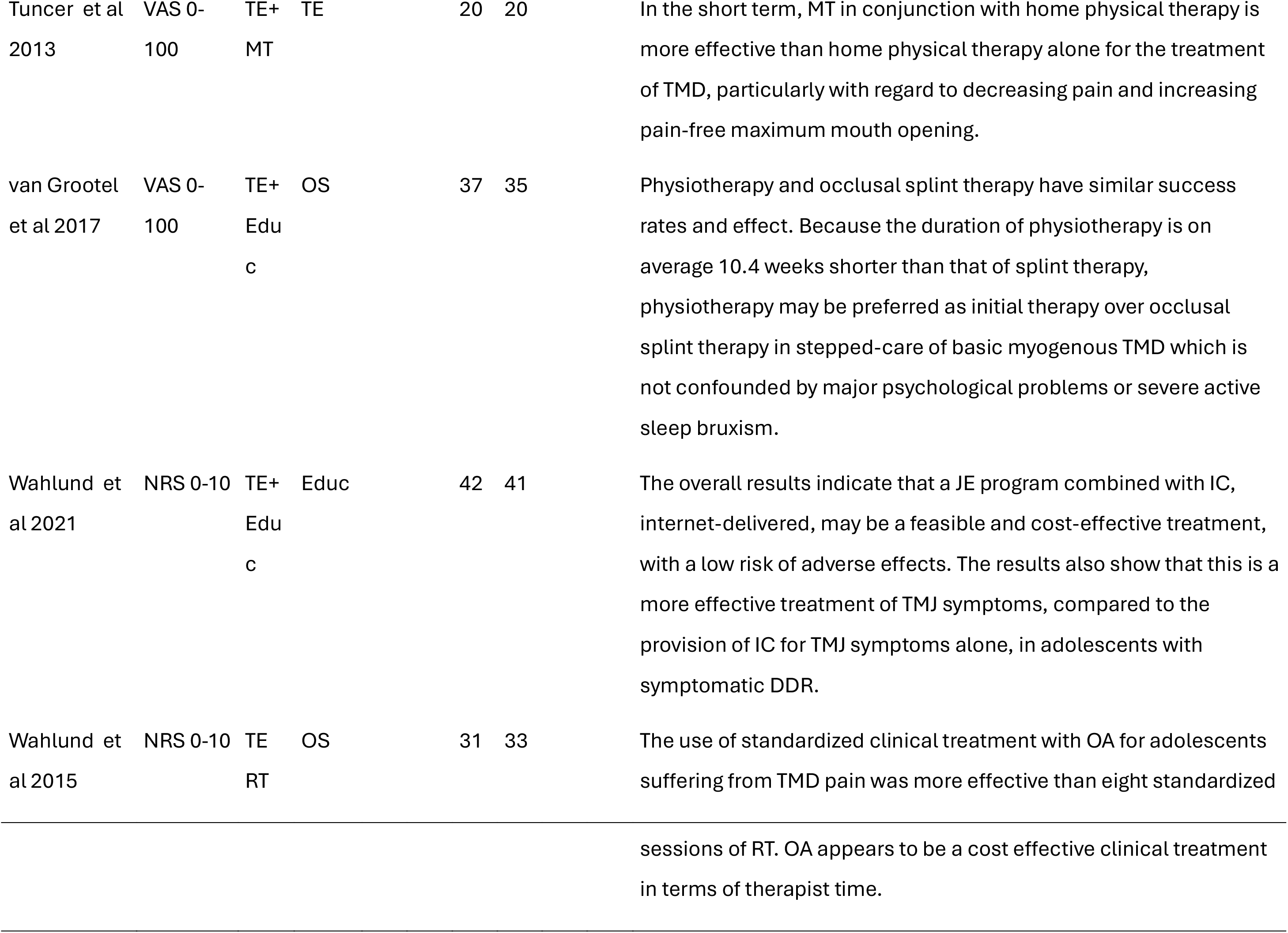
Characteristics of the included studies extraction. Abbreviations: Exp: experimental group, Ctr: control group, VAS: Visual Analog Scale, TE: Therapeutic Exercise, Plac: Placebo, Educ: Education, IPT: Instrumental Physical Therapy, OS: Occlusal Splint, NPRS: Numerical Pain Rating Scale, Dry N: Dry Needling, NSAID: Non-steroidal Anti Inflammatory Drugs, MT: Manual Therapy, KT: Kinesio Taping, NRS: Numerical Rating Scale

### Methodological quality

The methodological quality of the included studies was evaluated using the PEDro Score (Table 3). The mean score of the included articles was 6.32±1.55 points and the median was 7 points. The most observed criteria were randomization, comparability at baseline (100%), presence of results of between-group statistical comparisons (100%) and presence of both point measures and measures of variability (96%). Subjects were blinded to the treatment only in 12% of the studies [35,37,42], therapists were never blinded and assessors were blinded only in 68% of cases [26,27,29,30,32,34,35,36,37,39,41,42,43,44,46,47,48]. Allocation was concealed in sixteen of the included studies (64%) [25,27,30,31,32,34,35,36,37,41,42,43,44,46,48,49]. Only eight studies (32%) made an intention to treat analysis [26,27,30,31,32,33,35,48].

**Table 3.**
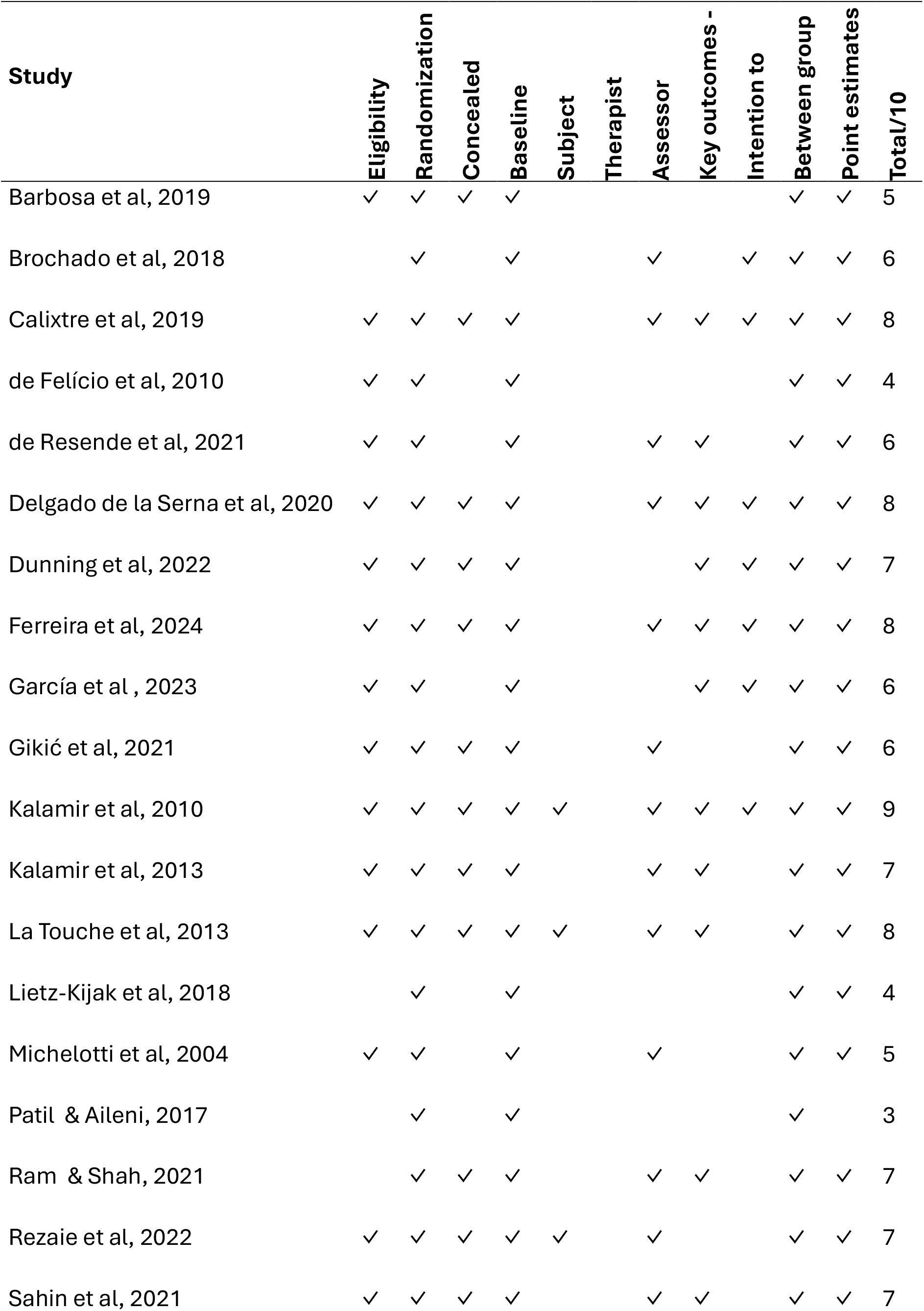
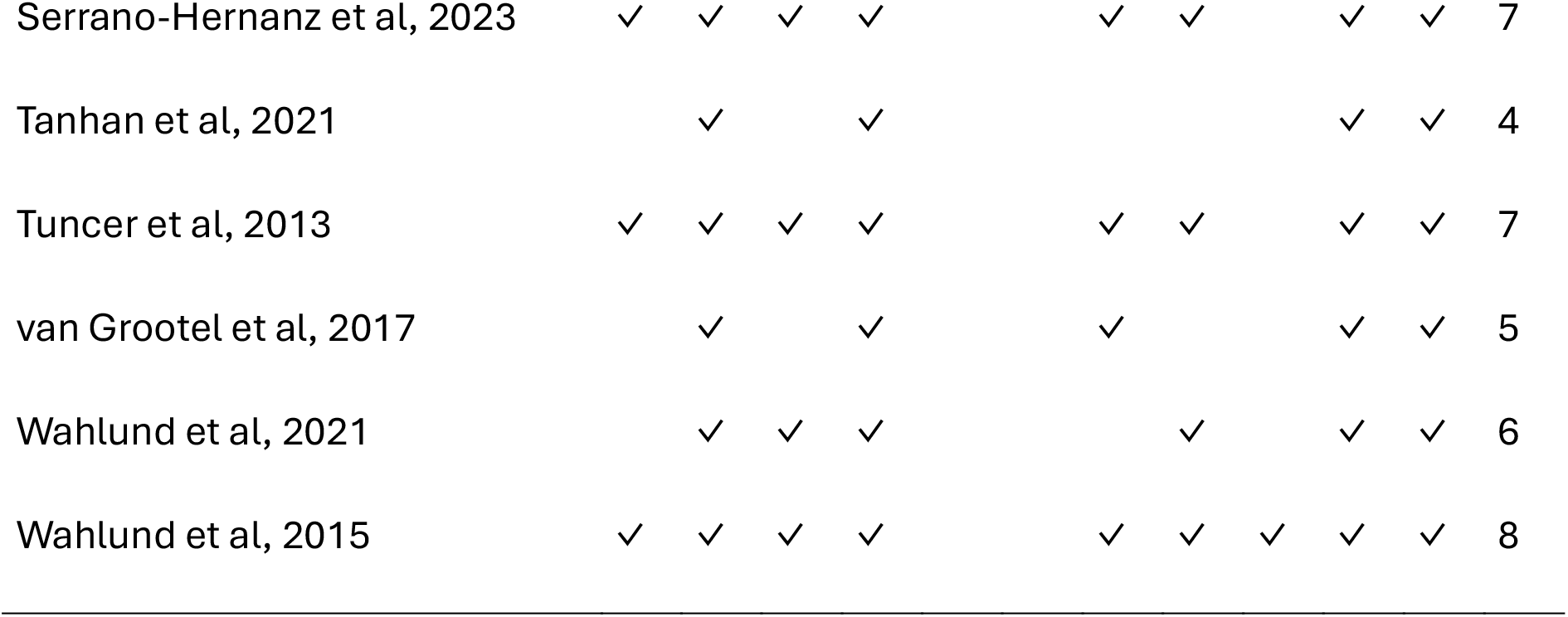
PEDro Score of the included articles.

### Summary of main findings

The synthesized evidence indicates that physical therapy interventions (therapeutic exercise and/or manual therapy) are more effective than no treatment or placebo for reducing pain in temporomandibular disorders (TMDs), although the certainty of evidence is influenced by significant heterogeneity across studies.

When compared to occlusal splints, most studies found no significant difference in effectiveness, positioning physical therapies as a valid alternative, often with a shorter treatment duration.

Comparisons between different active treatments (e.g., manual therapy vs. therapeutic exercise, or their combination) generally yielded mixed or similar results, suggesting multiple pathways can be effective.

The meta-analyses corroborate these trends: manual therapy demonstrated a moderate-to-large overall effect size (SMD -0.66), while therapeutic exercise showed a small, non-significant effect (SMD -0.16). The direct comparison favoured manual therapy but was not statistically significant (SMD -0.34). Significant heterogeneity was present, but publication bias was low. The forest plots (Figures 2, 3, and 4) provide detailed visualization of these pooled results.

**Figure 2.**
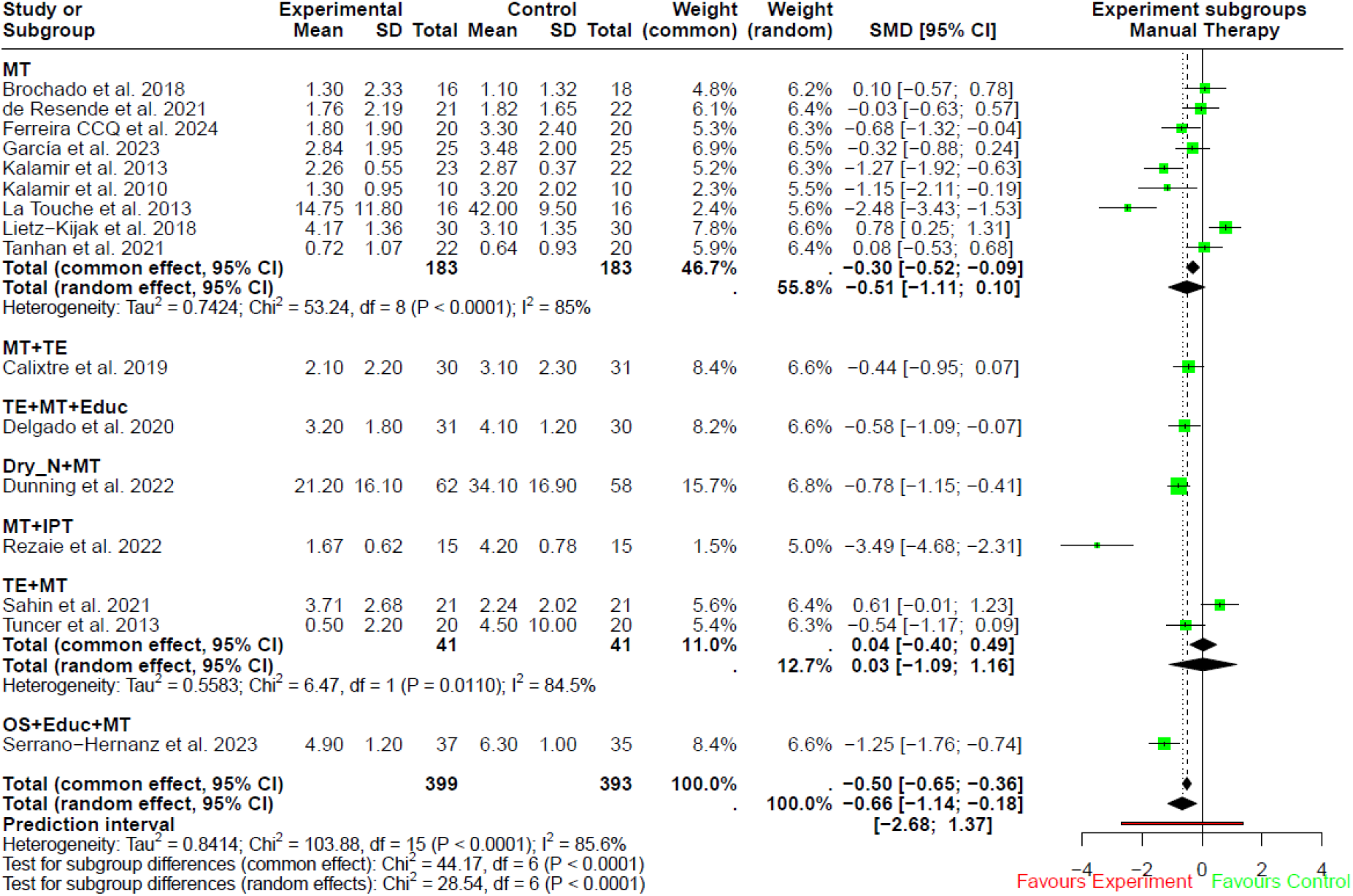
Forest plot illustrating the comparison between manual therapy vs other therapies to improve pain in temporomandibular disorders through a meta-analysis.

**Figure 3.** Forest plot illustrating the comparison between therapeutic exercise vs other therapies to improve pain in temporomandibular disorders through a meta-analysis.

**Figure 4.**
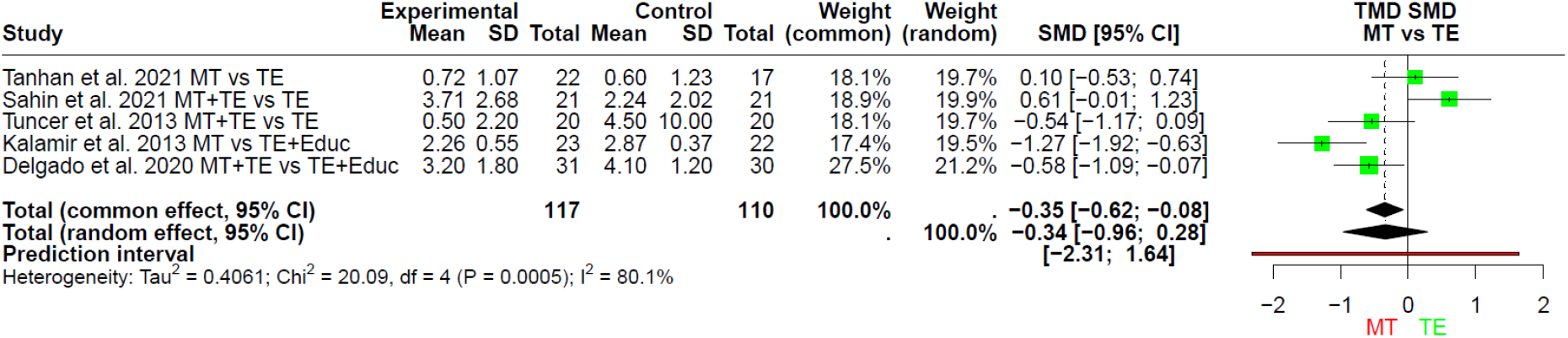
Forest plot illustrating the comparison between MT and TE to improve pain in TMDs through a meta-analysis.

The network meta-analysis was performed with seven multi-arm studies (five with three groups and two with four groups). Network of comparisons: nodes represent interventions; lines represent available direct comparisons between interventions, based on the included studies. Node size is proportional to the number of patients randomised per treatment, and line thickness is proportional to the number of studies for each direct comparison.

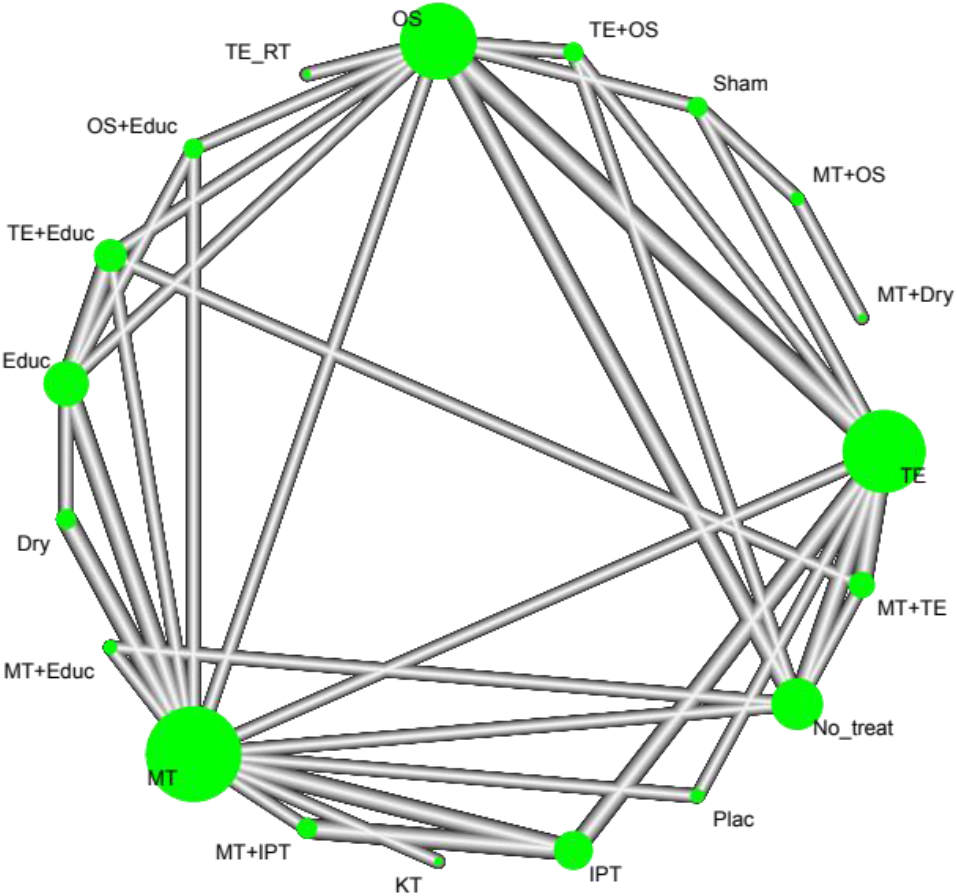

Upon checking the Q statistic, the studies by La Touche et al [34] (Q=42.47), Patil C Aileni [37] (Q=26.16), Brochado et al [24] (Q=13.81) and Rezaie et al [39] (Q=7.62) substantially contribute to the inconsistency of the network. Treatment estimates were obtained by comparing all other studies versus therapeutic exercise and were reported in the results section. The estimate of heterogeneity and inconsistency was tau^2=2.0612 I^2=85.1% IC [77.8%;90.0%] and the tests of heterogeneity within designs Q=5.34 and of inconsistency between designs Q=108.67 for a total Q of 114.01. Substantial heterogeneity indicates the use of random effects.

To confirm this choice, the total inconsistency of random effects was calculated [10]. The consistency under the assumption of a full design-by-treatment interaction random effects model was (between designs) Q of 40.30, an important but not significant reduction (p<0.0001). This could be because many estimates derive from indirect evidence. To evaluate the amount of indirect evidence and its propagation in the network, we produced a network graph. Finally, a heat map was drawn up showing that between fixed effects and random effects, the consistency among studies does not improve much (as suggested by the reduction in Q but not by its significance). Still, it increases the weight of the comparisons, supporting the estimate of random effects.

To further evaluate the consistency of the network, a forest plot was drawn up with direct and indirect estimates and their pooling. Comparisons of instrumental physical therapy vs manual therapy plus instrumental physical therapy, occlusal splint vs therapeutic exercise plus occlusal splint, occlusal splint vs therapeutic exercise plus education, occlusal splint vs therapeutic exercise, occlusal splint vs occlusal splint plus education, no treatment vs therapeutic exercise, manual therapy plus therapeutic exercise vs therapeutic exercise plus education, manual therapy vs therapeutic exercise show standardized mean differences with different signs but not significantly different. At the same time, manual therapy vs placebo and placebo vs therapeutic exercise show significant differences between direct and indirect effects. The treatment ranking was synthesized in the forest plot with confidence intervals using therapeutic exercise as the usual treatment (Figure 5). Manual therapy treatments combined with dry needling (SMD -13.22 IC[-20.85;-5.59] and combined with instrumental physical therapy standardized mean difference -2.85 IC[-5.55;-0.14]) show significant pain reductions compared to therapeutic exercise, while therapeutic exercise-only treatments show significantly better results compared to placebo groups.

**Figure 5.**
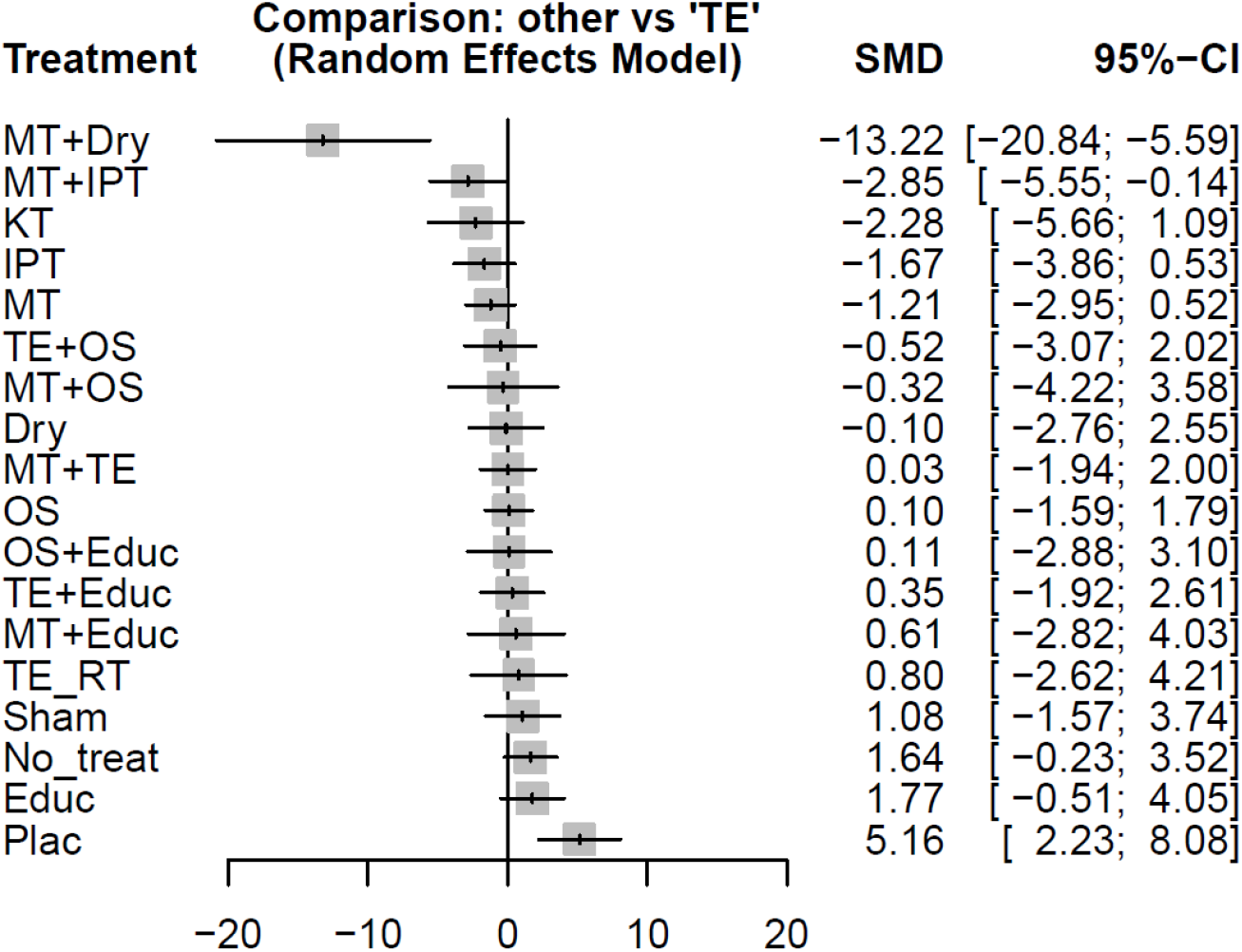
Ranking of various rehabilitative techniques compared to therapeutic exercise through a network meta-analysis.

## Discussion

This systematic review and network meta-analysis aimed to evaluate the effect of manual therapy and therapeutic exercise for reducing pain in individuals with temporomandibular disorders. The review included 25 randomized clinical trials with a total of 1,540 participants and analyzed both direct and indirect comparisons.

The results suggest that both manual therapy and therapeutic exercise are more effective than controls (including no treatment, placebo or education). Studies comparing these interventions with occlusal splints or instrumental therapies yielded mixed outcomes [26,28,31,34,40,41,42,44,45,47,48] with multimodal approaches (manual therapy + dry needling, manual therapy + instrumental physical therapy) often demonstrating superior pain reduction [31,42]. These findings align with a growing body of literature supporting combined interventions in rehabilitation.

Although both manual therapy and therapeutic exercise were effective, manual therapy showed slightly greater pain reduction in direct comparisons. However, heterogeneity across studies was considerable and confidence intervals were wide, indicating variability in treatment effects. This variability may reflect differences in study populations, intervention protocols, therapist expertise and outcome measurement tools.

Meta-analyses indicated that both manual therapy (SMD: -0.66; 95% CI: -1.18, -0.14) and therapeutic exercise (SMD: -0.16; 95% CI: -0.52, 0.21) reduce pain in TMDs, albeit with substantial heterogeneity (I² = 75-86%). Manual therapy demonstrated a clinically larger effect size than exercise, though their direct comparison was not statistically significant (SMD: -0.34; 95% CI: -0.96, 0.28). Heterogeneity decreased in subgroups comparing interventions to "no treatment." No significant publication bias was detected (p > 0.05).

The network meta-analysis included seven multi-arm studies and allowed for indirect comparisons of multiple treatments to determine the most effective therapeutic approach. The analysis revealed high heterogeneity (I²=86.7%) and significant inconsistency among study designs (Q=107.21, p<0.0001), justifying the use of a random-effects model.

The results indicated that the most effective strategies for pain reduction in temporomandibular disorders were manual therapy combined with dry needling (SMD - 13.23, 95% CI:-21.25;-5.4) and manual therapy combined with instrumental physical therapy (SMD -3.03, 95% CI:-5.90;-0.16). Conversely, placebo treatments and no treatment were significantly less effective. The treatment ranking analysis suggested that a multimodal approach, combining different rehabilitative strategies, may be the most effective therapeutic option for temporomandibular disorders.

Despite these findings, several limitations should be acknowledged for this review:

1. *Clinical heterogeneity*: variability in temporomandibular disorder subtypes, interventions and outcome assessment may limit the generalizability of results. Subgroup analyses based on diagnostic classification were not feasible. The time-related factor must be considered when establishing whether a therapy is effective or not, since some conditions have a shorter prognosis than others.
2. *Incomplete description of interventions*: many included studies lacked detailed reporting of the techniques used, limiting reproducibility and clinical translation. Also different treatment duration in the included studies was a limitation of this review.
3. *Risk of bias*: methodological quality varied widely. Two of the studies in which the control group gained a better result than the intervention group were characterized by a very low methodological quality (respectively 4 points for the study by Lietz-Kijak et al [38] and 3 points for the one by Patil et al [40]), so their results must be considered carefully; moreover, both of these studies showed an improvement in pain intensity also for the intervention group, even if significantly lower than the other group.
4. *Focus on pain only*: this review focused exclusively on pain outcomes without evaluating functional improvements or quality of life, which are important for holistic management of temporomandibular disorders: when interpreting the results of this study, it is important to consider the extreme subjectivity of pain, which is hard to quantify in a certain way. In a new era for physical therapy in which active coping strategies and physical activity are often preferred to passive strategies, it is reasonable to believe that other more functional outcomes would have better improved for different strategies. This element should be further investigated in future research.
5. *Lack of safety data*: the present review did not investigate the possible side effects of the proposed rehabilitative approaches. Even if the quantitative analysis showed a substantial effect of the conservative treatment with physical therapy, we cannot affirm that it is the safer one.

When selecting the inclusion criteria, the authors decided to exclude every comparison with invasive treatments such as surgery, mainly because it is not a conservative approach and physical therapists cannot administer it. During the study selection process, some interesting articles on this topic emerged: above all, there is the one by De Almeida et al [50], which compared a set of mandibular exercises with an arthrocentesis with sodium hyaluronate injection. This study highlighted that there are no significant differences between groups in post-treatment analysis, so the conservative approach may probably be chosen as the first one, even in this case.

This systematic review and network meta-analysis supports the effect of manual therapy and therapeutic exercise in reducing pain in individuals with temporomandibular disorders. These interventions should be considered first-line conservative treatments. Among all strategies, manual therapy combined with dry needling was identified as the most effective approach for pain relief, followed by manual therapy combined with instrumental physical therapy. Placebo, no treatment and education alone were the least effective. Given the variability in intervention protocols and study quality, further research is necessary to confirm these findings and improve clinical guidelines for temporomandibular disorders management.

We found that a multimodal treatment is the best therapeutic option to treat painful disorders of the temporomandibular joint.

### Clinical messages

Manual therapy and therapeutic exercise represent effective, reversible, and non-invasive strategies for the management of temporomandibular disorders. Given their analgesic efficacy and safety profile, these interventions should be prioritized as first-line treatments before the initiation of pharmacological regimens, occlusal appliances, or invasive procedures.

Evidence from randomized controlled trials indicates that both manual therapy and therapeutic exercise significantly reduce pain compared with placebo, education, or no intervention. Manual therapy demonstrated a trend toward greater analgesic effect sizes, although between-study heterogeneity and wide confidence intervals limit definitive conclusions regarding superiority.

Network meta-analysis highlights that multimodal rehabilitation strategies—particularly manual therapy combined with dry needling or adjunctive instrumental modalities— achieve superior pain reduction compared with unimodal approaches. These findings support the clinical adoption of integrative physiotherapeutic protocols to optimize outcomes in patients with temporomandibular disorders.

Substantial clinical and methodological heterogeneity, insufficient reporting of intervention protocols, and a narrow focus on pain outcomes restrict the generalizability of current evidence. Future high-quality randomized controlled trials should incorporate standardized intervention descriptions, functional and quality-of-life outcomes, and safety data to strengthen clinical guidelines for temporomandibular disorders management.

## Conflict of interest

All the authors declare that they had no conflicts of interest or funding sources during the elaboration of this manuscript.

**Clinical Trial Protocol number Prospero CRD42022347010**

## Acknowledgements

We thank Dr Leonardo Pellicciari and Dr Matteo Paci for valuable discussions and review.

## Data Availability Statement

Data available on request: the data presented in this study are available on request from the corresponding author.

